# Prevalance of Carbapenemase Production Among Clinical Gram-Negative Isolates in Bangladesh: A Comprehensive Study Using Disc Potentiation Test (DPT) Method

**DOI:** 10.1101/2024.12.30.24319797

**Authors:** Sumon Kumar Das, Nikhat Ara, Suma Mita Biswas, Afzal Sheikh

## Abstract

**Background:** The prompt identification of carbapenemases, particularly *Klebsiella pneumoniae* carbapenemases (KPC) and Metallo-β-lactamase (MBL) produced by *Klebsiella pneumoniae* and *Enterobacteriaceae*, is imperative in stemming the spread of pathogens carrying these antibiotic-resistant attributes. This retrospective study seeks to delve into the clinical and microbiological characteristics of patients affected by gram-negative bacteria producing KPC and MBL, shedding light on their profiles and impacts.

**Methods:** We analyzed 147 clinical isolates from tertiary care hospitals in Dhaka, Bangladesh. Bacterial identification and Minimum inhibitory concentration (MIC) analysis were performed using conventional culture and VITEK**®**2 compact system. Carbapenem non-susceptibility (Cns) was defined as a minimum inhibitory concentration (MIC) of ≥1 µg/ml for *Enterobacteriaceae* while *Pseudomonas aeruginosa* and *Acinetobacter baumannii* bacterial isolates Imipenem non-susceptibility was defined as an MIC of ≥2 µg/ml. Detection of KPC and MBL-producing isolates was performed using the disk potentiation test (DPT) method.

**Results:** All 147 clinical bacterial isolates demonstrated resistance to Imipenem (MIC 1 -≥16 µg/ml), with *Klebsiella pneumoniae* carbapenemase (KPC) detected in 26.53 % (39 out of 147) isolates and metallo-beta-lactamases (MBL) in 44.89% (66 out of 147) isolates. Carbapenem resistance was notably prevalent among older male patients of the “> 60” age group, with the ICU harboring the highest number of resistant isolates, primarily associated with *Acinetobacter baumannii* followed by *Klebsiella pneumoniae*.

**Conclusion:** Our findings unveiled a concerning prevalence of carbapenem resistance among clinical bacterial isolates, with notable proportions of KPC and MBL producers. The findings underscore the urgent need for effective surveillance and infection control measures to mitigate the spread of multidrug-resistant pathogens in healthcare settings.

## Introduction

Infections caused by drug resistant gram-negative *Enterobacteriaceae*, notably *Klebsiella pneumoniae* producing *Klebsiella Pneumoniae* Carbapenemases (KPCs), pose significant global concerns. These organisms cause formidable challenges to clinicians due to limited antibiotic options, resulting in unfavorable clinical outcomes. Carbapenems serve as the primary treatment for severe infections induced by extended-spectrum beta-lactamases (ESBLs), given the escalating incidence of fluoroquinolone resistance among *Enterobacteriaceae*. As a result, carbapenems are being used more frequently in clinical settings to treat these infections (1,2). *Klebsiella pneumoniae* Carbapenemase (KPC) is not the sole carbapenemase identified in *K. pneumoniae*; there are also reports suggesting the existence of metallo-beta-lactamases (MBL) with the ability to hydrolyze carbapenems (3,4). The horizontal transmission of carbapenemase genes, facilitated by mobile genetic elements such as self-transmissible plasmids or transposons, is a significant concern in antibiotic resistance. These elements not only carry carbapenemase genes but also harbor additional resistance determinants, leading to multidrug resistance. As a consequence, bacteria become resistant to multiple groups of antibiotics, often rendering them impervious to all available treatment options (5,6).

The management of infections caused by carbapenemase-producing *Enterobacteriaceae* has become exceedingly difficult owing to their profound resistance against a broad spectrum of antibiotics. Compounding the challenge is the inherent nature of *Enterobacteriaceae* as members of the intestinal flora, facilitating their rapid spread and resilient persistence. Furthermore, the rampant utilization of antibiotics in both human medicine and in the food chain, coupled with their subsequent release into the environment, serves to expedite the selection and horizontal transfer of antibiotic-resistant genes or plasmids within bacterial populations (2,7). As MBLs are predominantly encoded within transposons and/or integrons, they possess a high propensity for dissemination to other enterobacterial strains (7–9).

The primary reservoir for these bacteria is believed to be hospitals and other healthcare facilities, where instances of severe bacterial infections are prevalent and antibiotic usage is extensive (10,11). Numerous studies worldwide have investigated and assessed the efficacy of simple phenotypic tests for the specific identification of carbapenemase-producing strains, with varying degrees of endemicity observed based on the type of enzyme (12).

Various methods have been developed for the detection of carbapenemases, including the Modified Hodge Test (MHT) (13), the Carba NP (14) test and its derivatives, the modified carbapenem inactivation method (mCIM) (15), as well as antibody-and PCR-based approaches (16,17). However, traditional forms of these methods lack specificity regarding the type of carbapenemase produced and cannot distinguish between *Klebsiella pneumoniae* carbapenemase (KPC) and metallo-beta-lactamase (MBL) carbapenemase. Despite the ability of antibody-and PCR-based methods to differentiate between KPC and MBL, they cannot discern carbapenem-resistant *Enterobacteriaceae* (CRE) from carbapenem-susceptible *Enterobacteriaceae* (CSE) isolates, particularly in cases where antimicrobial susceptibility is unknown. Furthermore, these methods are costlier and less accessible in local healthcare settings compared to traditional detection methods, thereby posing limitations in meeting clinical demands effectively.

This study investigates the efficacy of the Disk Potentiation Test (DPT) for rapid and accurate identification of KPC and MBL carbapenemases, significant resistance determinants commonly found in *Enterobacteriaceae* isolates. By employing the DPT method, healthcare professionals can effectively identify KPC and MBL enzyme from CRE, aiding in the selection of appropriate antibiotics for patients with carbapenemase-producing CRE infections. This approach aims to mitigate the spread of carbapenemase-producing bacteria within healthcare settings.

## Materials & Methods

### Clinical isolates

From January to June 2020, in a six-month period, a total of 147 gram-negative bacterial isolates were collected from various units of Evercare Hospital, previously called Apollo Hospital, Dhaka, were used for this study. During routine laboratory procedures, all showing resistance to Imipenem. Carbapenem non-susceptibility (Cns) was defined as a minimum inhibitory concentration (MIC) of ≥1 µg/ml for *Enterobacteriaceae* while *Pseudomonas aeruginosa* and *Acinetobacter baumannii* bacterial isolates Imipenem non-susceptibility was measured as an MIC of ≥2 µg/ml using the VITEK® 2 compact system. Following this, all Imipenem-resistant gram-negative bacteria were screened for KPC and MBL using the disc potentiation test (DPT) method (18–20). The isolates were obtained from outpatients (OPD), inpatients (IPD), and intensive care unit (ICU) patients at Evercare Hospital in Dhaka. The Microbiology & Immunology laboratory conducted phenotypic testing for KPC and MBL detection using the DPT method, which has demonstrated higher detection sensitivity in our previous reports (21).

All experimental methods were carried out in accordance with guidelines and regulations approved by the “Research and Ethical Practice Committee” of Evercare Hospital, Dhaka, Bangladesh. Oral informed consent was obtained from the study subjects/patients or from their legal guardians and was approved by the “Research and Ethical Practice Committee” at Evercare Hospital, Dhaka, Bangladesh and the data were analyzed anonymously.

### Phenotypic confirmation of Disc potentiation test (DPT) for KPC detection

The presence of KPC in *K. pneumoniae* and *Enterobacteriaceae* isolates was determined using boronic acid disk tests (18). A stock solution of phenylboronic acid (benzeneboronic acid) from Sigma-Aldrich, Steinheim, Germany, was prepared by dissolving 120 mg of phenylboronic acid in 3 ml of dimethyl sulfoxide. Then, 3 ml of sterile distilled water was added to this solution to reach a final concentration of 20 g/L. From this solution, 20 μl (containing 400 μg of phenylboronic acid) was dispensed onto commercially available Imipenem disks. Before application, the disks were dried and employed within 60 minutes (18,22). The test relies on comparing the zone diameters produced by discs containing Imipenem (IPM) with or without phenylboronic acid (PBA). A difference in zone diameter of >5 mm between the IPM and IPM + PBA discs was considered a positive result (22–23).

### The phenotypic confirmation of metallo-beta-lactamase (MBL) detection through DPT method

The process involved the utilization of 0.5 M EDTA to inhibit MBL activity, as detailed by Yong et al. (20). This assay hinges upon comparing the zones generated by discs containing Imipenem (IPM) with or without 0.5 M EDTA Galani et al. (19) adapted the test by employing two 10 µg IPM discs. Subsequently, 10 µl of 0.5 M EDTA was applied to one of the IPM discs. The plates were incubated overnight at 37°C. In the DPT with 0.5 M EDTA, a growth inhibitory zone diameter enlargement exceeding 7 mm around the IPM + 0.5 M EDTA disc compared to IPM alone indicated a positive result for MBLs (19).

### Statistical Analysis

For the statistical analyses, data were entered into an Excel spreadsheet and analyzed using the statistical software SPSS version 27.0 (IBM; Chicago, IL, USA) for Windows.

## Results

### Universal resistance to Imipenem was observed among all 147 bacterial isolates, particularly affecting individuals over 60 years,

A total of 147 bacterial strains isolated from various clinical specimens were subjected to antibiogram test and exhibited 100% resistance to Carbapenems (Imipenem). We also investigated the distribution of resistant and susceptible isolates among different age groups. Results represented that the number of resistant isolates distribution was significantly high in >60 age group compare to other age groups (Table 1). The percentage of bacterial isolates from male and female patient groups that showed 100% resistant to Imipenem were 78.2% and 21.8 % respectively (Fig 1a). The frequency of males and females affected by CRE in different unit of the hospital, the highest number of male patients affected carbapenem non-susceptible in ICU unit of the hospital (Fig 1b). The age-group “0 -15” and “>60” also represented the maximum resistant isolates from ICU hospital unit compare to IPD, OPD units (Fig 1c). In addition, the ICU unit displayed the highest prevalence of resistant isolates, with *Acinetobacter baumannii* being the most prominent, followed by *Klebsiella pneumoniae*, *Pseudomonas aeruginosa*, and *Providencia stuartii*, while *Acinetobacter baumannii* was most frequently found in ICU units, and *Klebsiella pneumoniae*, *Pseudomonas aeruginosa*, and *Providencia stuartii* were predominant in IPD units (Fig 1d).

**Fig 1.**
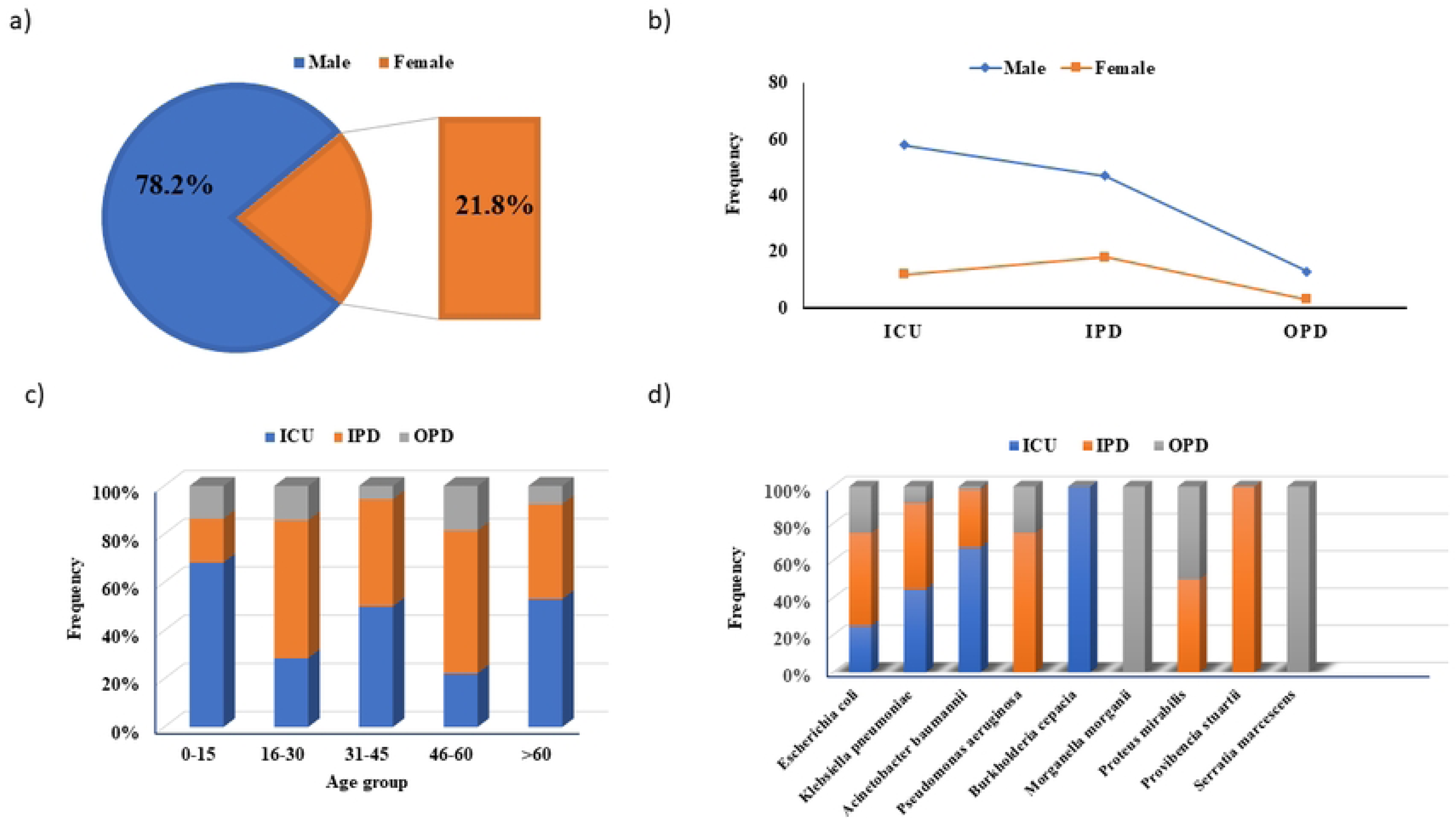
Resistance patterns of bacterial isolates across age and sex groups, depicting 100% resistance to carbapenems across all age categories, with the ">60" age group showing the highest number of resistant isolates. (a) Percentage of males and females affected by CRE. (b) Distribution of males and females affected by CRE in different hospital units. (c) Frequency of age-groups affected by CRE in different hospital units. (d) Frequency of clinically isolated carbapenem-resistant organisms (CRE) as etiological agents in clinical samples from various hospital units.

**Table 1.**
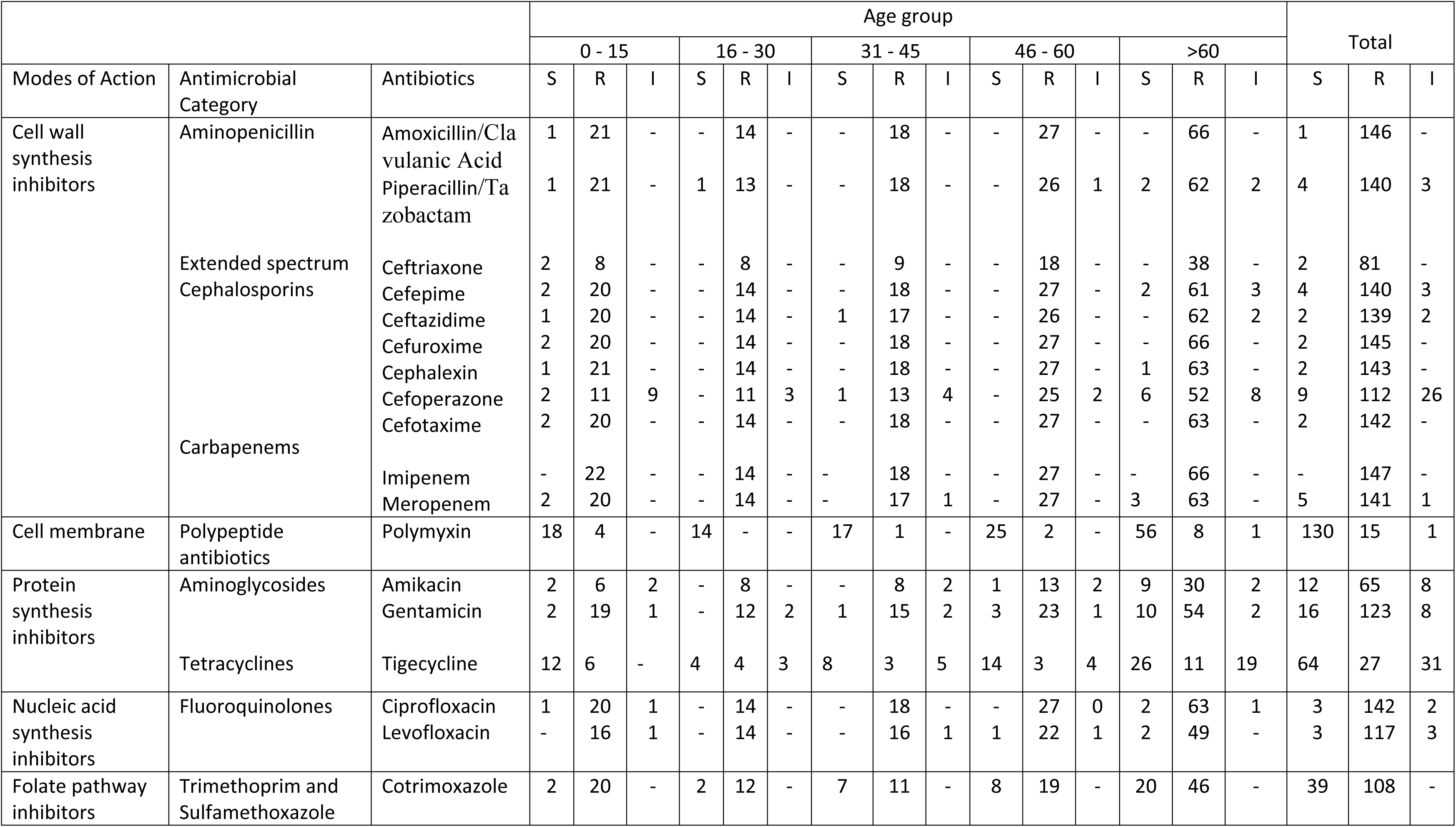
Represents the antibiotic susceptibility profiles of various gram-negative strains (n = 147) across different age groups, all of which exhibited 100% resistance to Imipenem. The susceptibility profiles were assessed for *Escherichia coli, Klebsiella pneumoniae, Acinetobacter baumannii, Pseudomonas aeruginosa, Burkholderia cepacia, Morganella morganii, Proteus mirabilis, Providencia stuartii, and Serratia marcescens* (n = 147) using conventional culture and the VITEK®2 compact system. Minimum inhibitory concentration (MIC) analysis was conducted in accordance with CLSI guidelines. Additionally, the distribution of resistant and susceptible isolates among age groups was examined, revealing the highest number of resistant isolates in the age group ">60".

### The “>60” age group exhibited the highest resistance, while a majority of gram-negative isolates displayed resistance to commonly used antimicrobial agents at MIC range of ≥16 µg/ml.

We also observed our observation of the distribution resistant bacterial isolates (n = 147) among age-groups data revealed that “>60” age-group showed the maximum number of resistant isolates (Table 2). We also analyzed MIC (Imipenem) -wise distribution spectrum of all gram-negative isolates. Individual bacterial isolates (n = 147) from MIC range 1 - 15, 16 - >16 were treated against commonly used antimicrobial agents*. Escherichia coli, Klebsiella pneumoniae, Acinetobacter baumannii, Pseudomonas aeruginosa, Proteus mirabilis, Serratia marcescens, Morganella morganii, Providencia stuartii, Burkholderia cepacia* showed resistant to antimicrobial agents. Highest number of resistances was showed at MIC ≥16 µg/ml (n = 126) (Table 2).

**Table 2.**
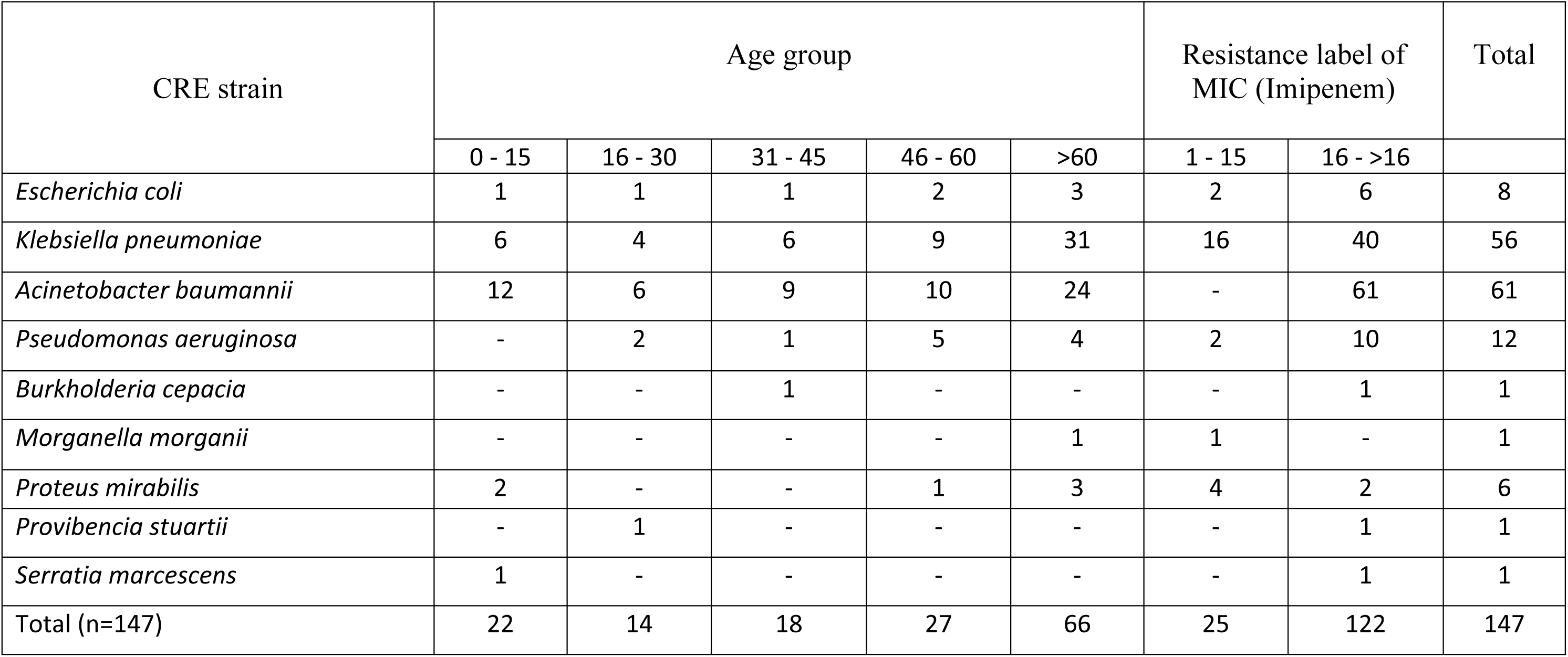
Age-group distribution spectrum of various isolates and their MIC-based resistance level to Imipenem. A total of 147 bacterial isolates were subjected to antimicrobial susceptibility testing across different age groups, ranging from 0 to >60 years. *Escherichia coli, Klebsiella pneumoniae, Acinetobacter baumannii, Pseudomonas aeruginosa, Burkholderia cepacia, Morganella morganii, Proteus mirabilis, Providencia stuartii, and Serratia marcescens* exhibited resistance to commonly used antimicrobial agents, with isolates from various MIC ranges showing similar resistance patterns.

#### Acinetobacter baumannii accounted for the highest detection of both KPC and MBL enzyme

Our study found that *Acinetobacter baumannii* strains had the highest occurrence of both KPC and MBL enzymes. Among the bacterial isolates, 36 (24.49%) exhibited both KPC and MBL enzymes, 78 (53.06%) showed no enzyme presence, 3 (2.04%) had the KPC enzyme, and 30 (20.40%) had the MBL enzyme. Specifically, *Acinetobacter baumannii* accounted for the highest detection with 30 strains, followed by *Klebsiella pneumoniae* with 3 strains, *Pseudomonas aeruginosa* with 2 strains, and *Escherichia coli* with 1 strain (Table 3).

**Table 3.** Crosstabulation analysis KPC and MBL enzymes profiles of various gram-negative strains (n=147). Interaction status of CRE strains to profile KPC and MBL drug-resistant enzymes among various gram-negative bacteria, including *Escherichia coli, Klebsiella pneumoniae, Acinetobacter baumannii, Pseudomonas aeruginosa, Burkholderia cepacia, Morganella morganii, Proteus mirabilis, Providencia stuartii, and Serratia marcescens* were examined.

| CRE strains | Types of drug resistant enzymes detected |  |  |  | Total |
| --- | --- | --- | --- | --- | --- |
|  | No enzyme | KPC and MBL<br>(both enzyme) | KPC enzyme | MBL enzyme |  |
| *Escherichia coli* | 2 | 1 | 1 | 4 | 8 |
| *Klebsiella pneumoniae* | 39 | 3 | 0 | 14 | 56 |
| *Acinetobacter baumannii* | 24 | 30 | 2 | 5 | 61 |
| *Pseudomonas aeruginosa* | 6 | 2 | 0 | 4 | 12 |
| *Burkholderia cepacia* | 1 | 0 | 0 | 0 | 1 |
| *Morganella morganii* | 1 | 0 | 0 | 0 | 1 |
| *Proteus mirabilis* | 5 | 0 | 0 | 1 | 6 |
| *Providencia stuartii* | 0 | 0 | 0 | 1 | 1 |
| *Serratia marcescens* | 0 | 0 | 0 | 1 | 1 |
| Total (n=147) | 78 (53.06%) | 36 (24.49%) | 3 (2.04%) | 30 (20.40%) | 147 |

### Detection of KPC and MBL Enzymes in CRE Isolates via DPT Method Shows High Prevalence in ICU Patients

We investigated the prevalence and distribution of KPC and MBL carbapenemases among clinical isolates. Among the clinical isolates investigated, 26.53 % (39 out of 147) were carriers of the KPC enzyme, and were predominantly identified in the ICU (Fig 2a). The distribution isolates are found to be KPC positive for these isolates varied across different units, with the highest prevalence noted in the ICU (23 out of 69), followed by IPD (14 out of 62) and OPD (2 out of 16). Notably, the majority of KPC-positive isolates exhibited resistance to carbapenems, with a significant portion falling within the MIC range, ≥16 µg/ml (Fig 2b). *Acinetobacter baumannii* emerged as the predominant bacterial strain producing the KPC enzyme accounting for 32 out of 61 isolates (Fig 2c). In addition, we analyzed clinical samples using the DPT method to detect CRE bacterial isolates with both KPC and MBL enzymes. The distribution and interaction of these drug-resistant enzymes were profiled among various gram-negative strains. The highest number of samples with both KPC and MBL enzymes were found in tracheal aspirates (n = 13), followed by suction tips (n = 4), blood (n = 4), wound swabs (n = 4), urine (n = 4), endotracheal tube tips (n = 3), sputum (n = 2), and pus (n = 2) (Table 4, Fig 2d). On the other hand, among the 147 clinical isolates examined, 44.89% (66 out of 147) tested positive for MBL using the DPT test with 10 µl of 0.5 M EDTA, with the highest prevalence observed in the ICU (33 out of 69), followed by IPD (28 out of 62) and OPD (5 out of 16) (Fig 2a). The majority of MBL-positive isolates displayed carbapenem resistance, with the highest frequency observed (Fig 2b). *Acinetobacter baumannii* was identified as the predominant MBL-producing bacterial strain (Fig 2c). Additionally, the highest occurrences of bacterial isolates resistant to carbapenem carrying the MBL enzyme were found in tissue samples, followed by endotracheal tube tips, suction tips, blood, urine, tracheal aspirate, wound swab, pus, and sputum (Fig 2d). The majority of KPC and MBL both enzymes displayed ICU department n = 22, followed by IPD n = 12 and OPD n = 2 (Table 5).

**Fig 2.**
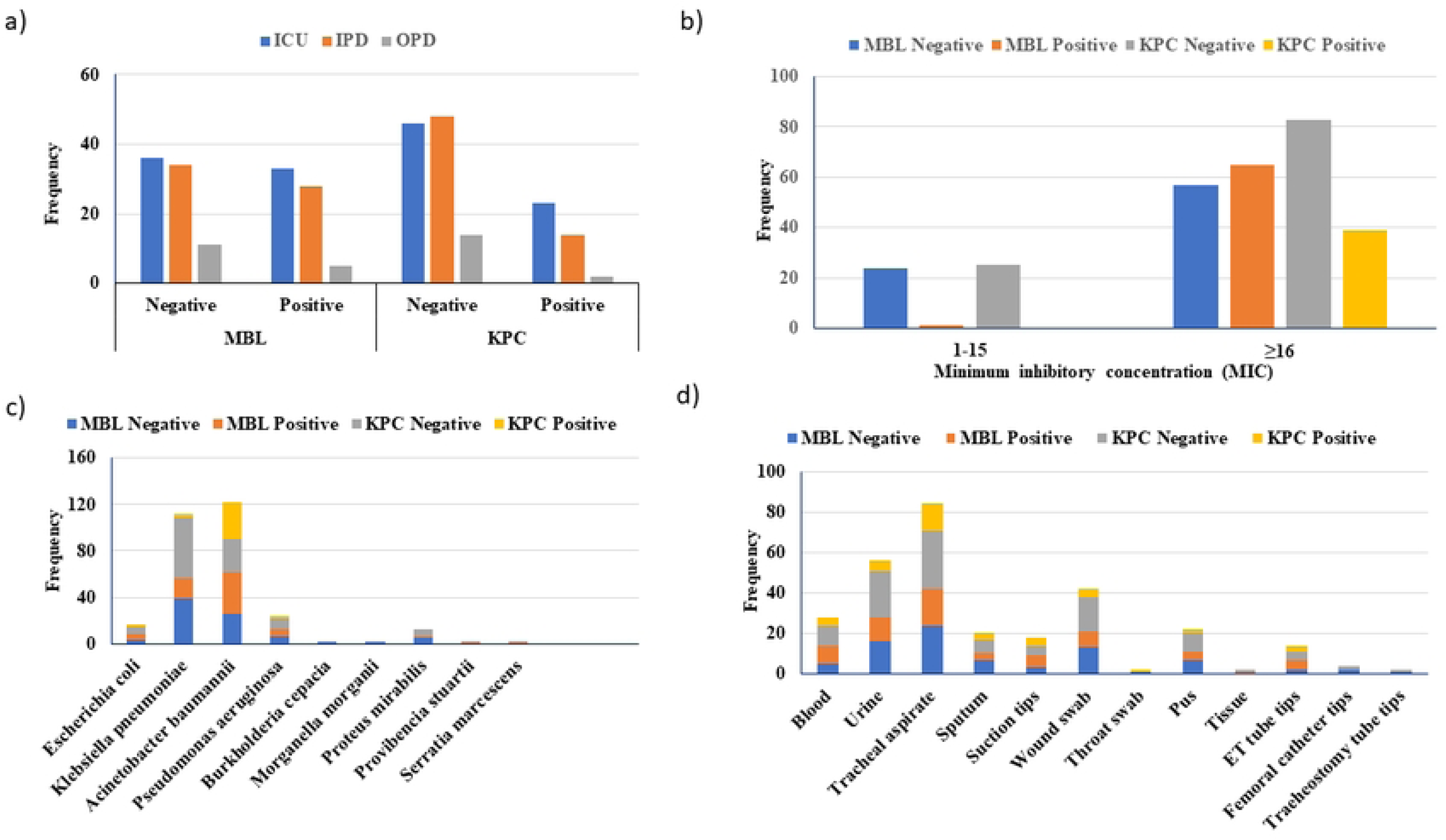
Distribution and screening outcomes of KPC and MBL enzymes in clinically isolated carbapenem non-susceptible pathogens:: (a) The count of KPC and MBL enzyme screenings using various inhibitors across different hospital units, (b) The incidence of KPC and MBL enzyme isolation based on MIC distribution, (c) The number of KPC and MBL enzyme screenings affected by clinically isolated etiological agents using different inhibitors, and (d) The frequency of KPC and MBL enzyme screenings across various clinical samples.

**Table 4.**
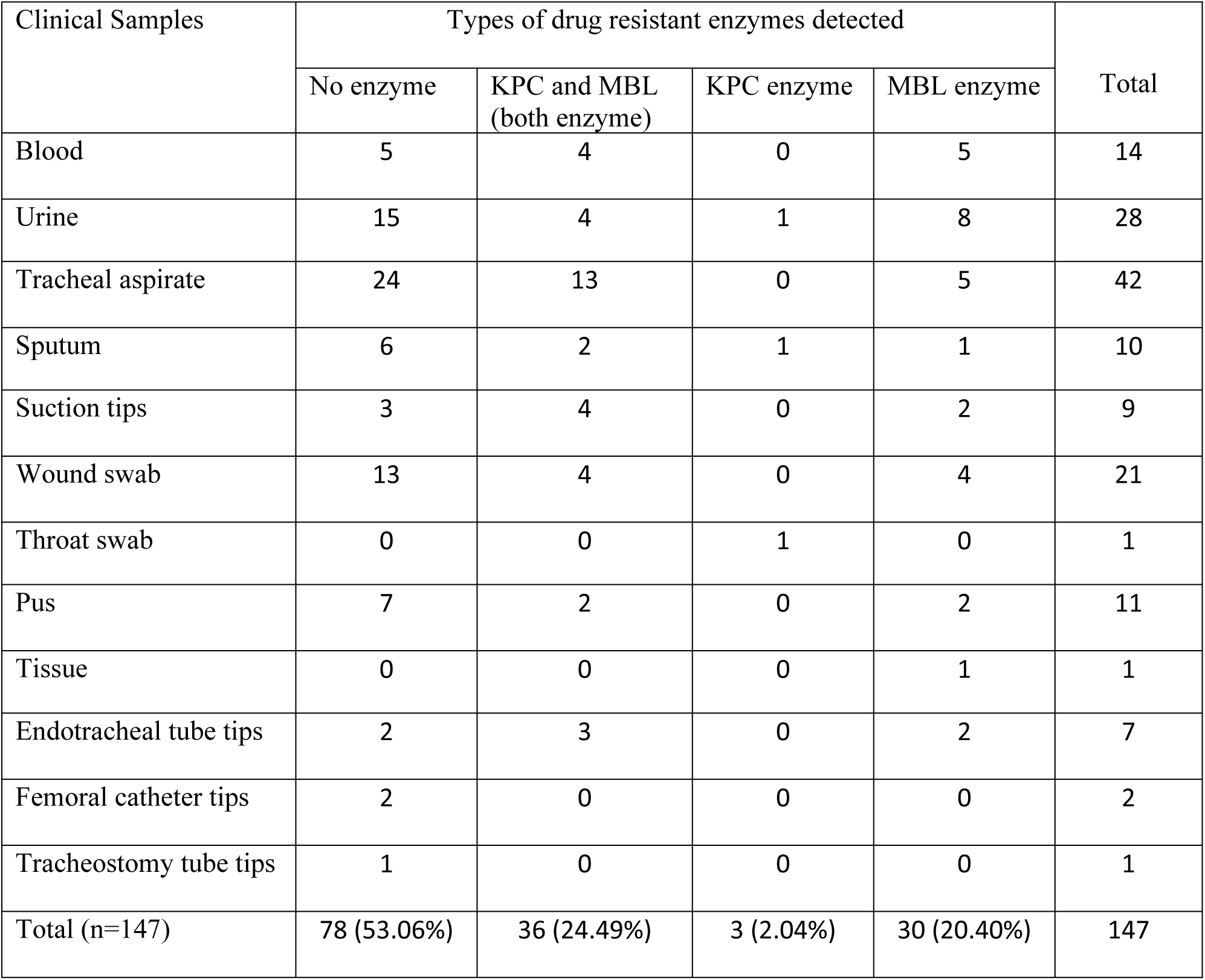
Crosstabulation analysis KPC and MBL enzymes profiles of various Clinical samples origin. In this analysis the distribution and interaction of clinical samples to profile KPC and MBL drug-resistant enzymes among various gram-negative strains, including *Escherichia coli, Klebsiella pneumoniae, Acinetobacter baumannii, Pseudomonas aeruginosa, Burkholderia cepacia, Morganella morganii, Proteus mirabilis, Providencia stuartii, and Serratia marcescens* were examined.

**Table 5.** Crosstabulation analysis KPC and MBL enzymes profiles of various unit of the hospital. In this analysis the distribution and interaction of ICU, IPD and OPD unit to profile KPC and MBL drug-resistant enzymes among various gram-negative strains, including *Escherichia coli, Klebsiella pneumoniae, Acinetobacter baumannii, Pseudomonas aeruginosa, Burkholderia cepacia, Morganella morganii, Proteus mirabilis, Providencia stuartii, and Serratia marcescens* were examined.

| Department | Types of drug resistant enzymes detected |  |  |  |  |
| --- | --- | --- | --- | --- | --- |
|  | No enzyme | KPC and MBL<br>(both enzyme) | KPC enzyme | MBL enzyme | Total |
| ICU | 35 | 22 | 1 | 11 | 69 |
| IPD | 32 | 12 | 2 | 16 | 62 |
| OPD | 11 | 2 | 0 | 3 | 16 |
| Total | 78 (53.06%) | 36 (24.49%) | 3 (2.04%) | 30 (20.40%) | 147 |

## Discussion

Our study provides valuable insights into the prevalence and distribution of carbapenemases, particularly KPC and MBL, among clinical isolates, shedding light on critical factors influencing resistance patterns and patient outcomes. A striking finding of our investigation is the universal resistance to Imipenem observed among all 147 bacterial isolates, underscoring the urgent need for effective antimicrobial stewardship and infection control measures. Notably, individuals aged over 60 years exhibited a significantly higher prevalence of resistance, highlighting the potential impact of age-related factors on susceptibility to carbapenem-resistant infections. (Table 1).

The resistance levels to carbapenems, aminopenicillins, extended-spectrum cephalosporins, aminoglycosides, tetracyclines, and polypeptide antibiotics were unexpectedly high, consistent with findings from prior studies. For instance, Smita Sood in a study India Jaiypur has reported 88.33% of the isolates were found to be MBL producers, 4 (6.66%) were found to be MBL and KPC co-producers phenotypic confirmation was made by combined disc method (24), although the number of clinical isolates was insufficient to draw definitive conclusions including the age and sex distribution of the resistant isolates. Additionally, Datta et al. reported a significant increase in carbapenem resistance from 2.5% in 2002 to 52% in 2009, along with rising resistance to cefotaxime (75 to 97%) and piperacillin/tazobactam (55-84%) among *Klebsiella pneumoniae* strains isolated from bloodstream infections in Delhi (25). Likewise, research conducted in southern India reported the rates of carbapenem resistance among hospital-acquired *Klebsiella pneumoniae* isolates: Meropenem 43.6%, Imipenem 32%, Ertapenem 20.3%, and Colistin 60% (26). More recently, a study conducted in Dhaka, Bangladesh, reported a 4.8% incidence of resistant against carbapenems (Meropenem and Imipenem) and all the carbapenem resistant isolates were found to be KPC positive however were detected using the modified Hodge test methods, typically requires more steps and interpretation, making it relatively more complex which has less accessibility in local hospital settings (27). This pervasive resistance underscores the urgent need for effective antimicrobial stewardship programs and infection control measures.

Gender disparities in antimicrobial resistance were observed, with a higher percentage of resistant isolates identified among male patients compared to females, especially in the ICU, highlighting the need for targeted surveillance. The ICU exhibited a significant concentration of carbapenem-resistant strains compared to OPD and IPD (Fig 1), particularly among patients over 60 years old, with *Klebsiella pneumoniae* was the most prevalent organism isolated followed by *Acinetobacter baumannii*, *Pseudomonas aeruginosa*, and others (Table 2).

We also investigated the prevalence and distribution of KPC and MBL carbapenemases among clinical isolates. The finding revealed that 26.5% of the isolates carried the KPC enzyme, with a higher prevalence in the ICU. A high-level resistance across gram-negative isolates were observed at MIC values above 16 µg/ml for Imipenem, with *Acinetobacter baumannii* emerged as the primary producer of the KPC enzyme. Additionally, 45% of the clinical isolates tested positive for MBL, predominantly in the ICU. Isolates resistant to carbapenems due to KPC and MBL enzyme exhibited distinct frequencies among clinical samples, with tissue samples, endotracheal tube tips, suction tips, blood, urine, tracheal aspirates, wound swabs, pus, and sputum specimens demonstrating varying levels of resistance (Fig 2). In our investigation of carbapenemase-producing isolates by specimen types, predominantly observed in tracheal aspirate followed by urine, blood, wound swab, and others. A dissimilarity in the trend was observed by a study in Bangalore, India, where most carbapenem-resistant *Klebsiella pneumoniae* isolates were obtained from urine samples (28). And, evidenced by studies conducted in the USA and India have reported a higher prevalence of *Klebsiella pneumoniae*, Carbapenem-resistant (CRKP) in respiratory or blood specimens from adult male patients admitted to the ICU (29).

Our study identified *Acinetobacter baumannii* as the predominant carrier of both KPC and MBL enzymes among the bacterial isolates and was accounted for the highest detection of both the enzymes, with 30 strains, followed by *Klebsiella pneumoniae*, *Pseudomonas aeruginosa*, and Escherichia coli (Table 3). Furthermore, our analysis using the Disc Potentiation Test (DPT) method revealed a significant prevalence of KPC and MBL enzymes in clinical samples. The distribution showed that tracheal aspirates had the highest number of samples with both enzymes, followed by suction tips, blood, wound swabs, urine, endotracheal tube tips, sputum, and pus (Table 4).

These findings highlight the urgent need for robust monitoring and control measures to manage the spread of multidrug-resistant organisms, particularly *Acinetobacter baumannii*, in healthcare settings. The high prevalence of KPC and MBL enzymes in tracheal aspirates suggests a significant infection and transmission risk, underscoring the importance of strict infection control practices and the development of novel therapeutic strategies. Effective detection and management strategies of these resistant strains is critical to mitigating their impact on public health.

## Data Availability

All data generated or analyzed during this study are included in this published article

## Declaration statement

### Availability of data and materials

All data generated or analyzed during this study are included in this published article.

## Acknowledgments

This work was partially supported by personal fund of Dr. NA in Evercare Hospital Dhaka, Bangladesh. All authors contributed to this research work from their responsibility to the society. We are thankful to our colleagues, and laboratory attendant Ilias Hossain who supported this research work. We are grateful to the patients for providing samples and for their participation in our research.

